# Effects of behavioral restrictions on COVID-19 spread

**DOI:** 10.1101/2022.08.08.22278490

**Authors:** Kenji Sasaki, Yoichi Ikeda, Takashi Nakano

**Affiliations:** Center for Infectious Disease Education and Research, Osaka University, Osaka 565-0871, Japan; Research Center for Nuclear Physics, Osaka University, Osaka 567-0047

**Keywords:** COVID-19, epidemic model, broken-link model, lockdown measure, Omicron variant

## Abstract

Several measures including behavioral restrictions for individuals have been taken to control the spread of COVID-19 all over the world. The aim of these measures is to prevent infected persons from contacting with susceptible persons. Since the behavioral restrictions for all citizens, such as the city-wide lockdown, are directly linked to stagnation of economic activities, the assessment of such measures is crucial. In order to evaluate the effects of behavioral restrictions, we employ the broken-link model to compare the situation of COVID-19 in Shanghai where the lockdown was implemented from March to June 2022 with it in Taiwan where a spread of COVID-19 was known to be well controlled so far. The result shows that the small link-connection probability is achieved by substantial isolation of infected person including the lockdown measures. Although the strict measures for behavioral restrictions are effective to reduce the total infected people, the daily confirmed cases follow the curve which is evaluated by the broken-link model. This result is considered as unavoidable infections for population.

## 1. Introduction

A novel coronavirus occurred in Wuhan in 2019 [1,2] was spread over the world. More than two years later, the virus is still mutating and causing infections around the world, with a cumulative total of 550 million infections and 6.3 million deaths.

Several measures have been taken to suppress the spread of COVID-19 all over the world. Basic idea is to prevent infected individuals from contacting with susceptible people. There are roughly two ways which are monitoring method and/or lockdown method to restrict citizen’s activity.

The monitoring method is a primitive but efficient strategy to pick up the person who is required a treatment or isolation. In this method, thorough contact tracing and quarantine of infected people are made as needed by collecting positional information of infected individuals. However, due to invasions of privacy, this method has been applied by limited countries, such as China [3,4], Taiwan [5–7] and South Korea [8,9].

The strongest measure to control a spread of infectious diseases is considered to be the (local) lockdown which forces one not to contact with the others in the community regardless of one’s health condition. The lockdown method has been implemented in many countries and is believed to have reduced the size of the spread of infectious diseases and avoided the collapse of the medical system caused by the rapid increase in the number of infected patients. However, it is difficult to sustain the lockdown on a long-term since it has caused significant damage to people’s daily lives and economic activities.

China is still trying to achieve “zero COVID” by the monitoring method with the help of Information Technologies. Indeed, it is credited with suppressing the numbers of COVID-19 patients from 2020 to 2021 after the outbreak in Wuhan, reducing the cases to a significantly lower level. However, after two years from outbreak in Wuhan, the situation has changed by emergence of Omicron variant which has a strong infectivity via not only droplet but also aerosol transmission and can easily breaks through immunity gained by vaccination. Due to the surge of COVID-19 by Omicron variants, massive outbreaks have occurred in many parts of the world, and the lockdown method has been made to subside it in China.

There have been some evaluations of the extent to which the local lockdown or behavioral restrictions of infected persons have reduced the spread of infectious diseases [10–13]. This is because the counterfactual assumption based on the mean field SIR model over the population in which the cases continue to increase by the basic reproduction number, *R*_0_, for every infected persons in the infection period till the hard immunity is achieved. Consequently it would lead to the conclusion that any measures are always effective if the daily confirmed cases tend to be reduced if it is not considering the transmission networks between individuals. The goal of this paper is to avoid such problems and to estimate the effects of the measures to restrict social activity by using the broken-link model.

## 2. Methods

We briefly review the broken-link model [14,15] which is proposed as a new compartment model with the consideration of unconnected inflectional transmission link. The cumulative number of cases in this model is described by the Gompertz function with three parameters as the cumulative number of infected people *N*_∞_, the connection probability of transmission links *k*, and the basic reproduction number *R*_0_ = −*a*/ ln *k* with constant *a*. These parameters are determined by fitting procedure to the reported data.

In SIR model which is traditional compartment model first developed by Kermack and McKendrick in 1927 [16], the basic reproduction number *R*_0_ is the averaged number of cases directly generated by one case in populations where all individuals are susceptible to infections. The *R*_0_ is composed by several factors including the duration of infectivity of affected individual, the infectiousness of the virus, and the contact frequency of infected people in the population, so that it is determined on the regional basis. The *R*_0_ in the broken-link model is defined in the similar sense and can be compared with that in SIR model.

The surge of infectious disease analysed the broken-link model always subsides without any measure because the infected people is quarantined by oneself if PCR testing result is positive either symptomatic or asymptomatic, which leads to the *k* less than 1. The small *k* is favorable for suppression of infectious diseases and is achieved by a strong quarantine system under the monitoring method.

In this paper, the effects of behavioral restriction including lockdown are evaluated by comparing the sizes and link connection probabilities in two surges in Taiwan where the COVID-19 was well controlled without lockdown and one surge in Shanghai where the lockdown was implemented for suppressing the Omicron surge.

## 3. Results

### 3.1. The Alpha surge in Taiwan

From the beginning of May to the mid July 2021, the surge of COVID-19 by Alpha variant of the novel coronavirus occurred in Taiwan. Though measures such as school blockages were taken, the lockdown measure was not taken. But “zero COVID” policy with redthe monitoring method took place in this period. As a result of Alpha surge, about 13 thousands people were infected in Taiwan.

The epicurve whose numbers are reported by the Center for Systems Science and Engineering (CSSE) at Johns Hopkins University [17] is fitted by reda single Gompertz curve called a *wave*. Both the numbers of the daily confirmed cases shown in Fig. 1 (a) and the K-values in Fig. 1 (b) are well reproduced with the fit parameters listed in Table 1. It is interesting to compare the *k* in the Alpha surge with it in the first surge in Taiwan from 20th March to 10th April 2020. The link connection probability in the Alpha surge is larger than *k* ≃ 0.85 (*K*^′^ ≃ 0.0524) in the first surge [14].

**Figure 1.**
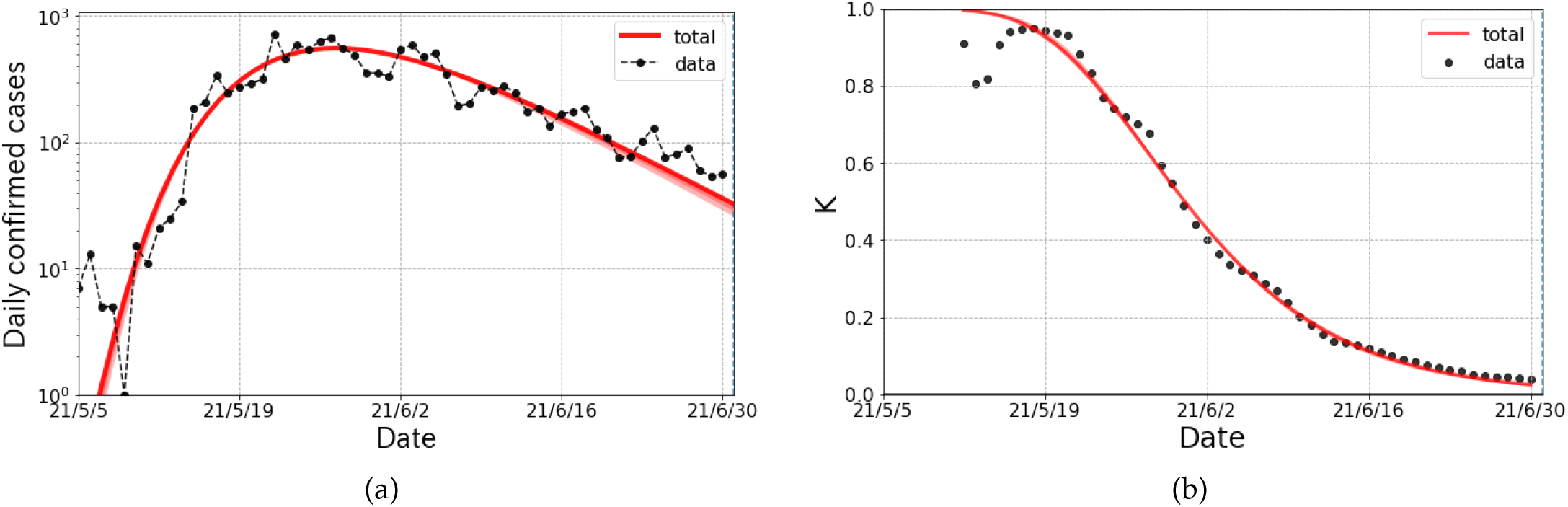
The logarithmic plot of the epicurve for the Alpha surge of COVID-19 in Taiwan from May to July 2021. (a) The he daily confirmed cases and fit results (solid curve). The fit is performed with a single partial wave denoted by dashed lines. (b) The observed data and fit result of the K-value. The red bands stand for systematic errors for the choice of fit range of the partial wave.

**Table 1.** The parameters of Gompertz curves for the Alpha surge in Shanghai. The *N*∞, *R*_0_ and *k* are the cumulative number of infected people, the basic reproduction number and connected probability of transmission links, respectively. The “Shift” stands for the onset of Gompertz curves from the reference date (5th May 2021).

| partial wave | $N_\infty$ | $R_0$ | $k$ | Shift |
| --- | --- | --- | --- | --- |
| 1st wave | 14k | 4.92 | 0.895 | 7.0 |

This leads that, under the same policy, the *k* is more or less the same in magnitude. Effects of changing from *k* = 0.85 to *k* = 0.90 is about 5 times enhancement in the number of daily confirmed cases at a perk.

### 3.2. The Omicron surge in Taiwan

For the period of the Omicron surge in Taiwan, they changed the policy of “zero COVID” to “pandemic management” policy [18–20]. After the mitigation of measures, the daily confirmed cases markedly increased by proliferation of BA.1.1 and BA.2 [21] in terms of PANGO (Phylogenetic Assignment of Named Global Outbreak) Lineages [22]. As shown in Fig. 2, the Omicron surge in Taiwan is decomposed into two waves whose onsets are synchronized to the emergence of new variants of coronavirus. From Tab. 2, the size of the first wave is quite small comparing with the size of second wave. Thus we mainly discuss the second wave though the first wave is indispensable to reproduce the epicurve.

**Figure 2.**
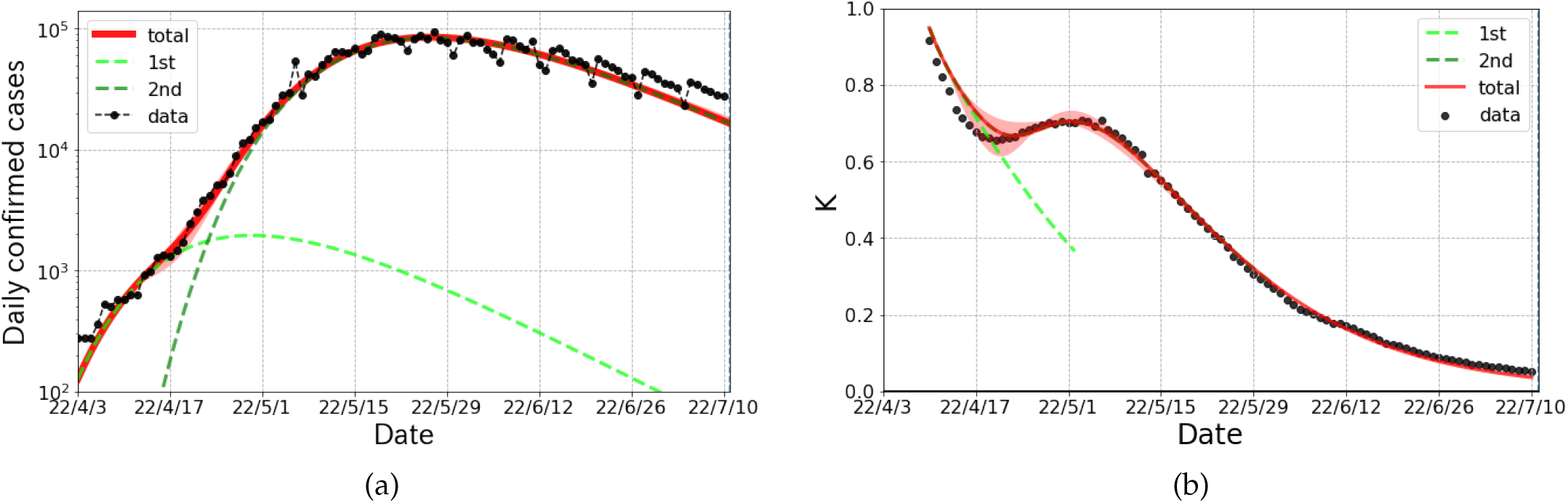
The logarithmic plot of the epicurve for the Omicron surge of COVID-19 in Taiwan from April to July 2022. (a) The daily confirmed cases and fit results (solid curve). The fit is performed with two partial waves denoted by dashed lines. (b) The observed data and fit results of the K-value. The red bands stand for systematic errors for the choice of fit range of the first and second partial wave.

**Table 2.** The parameters of Gompertz curves in the Omicron surge in Taiwan. The definition of the parameters is the same as in Table 1, but the reference date is 3rd April 2022.

| partial wave | $N_{\infty}$ | $R_0$ | $k$ | Shift |
| --- | --- | --- | --- | --- |
| 1st wave | 110k | 7.00 | 0.943 | -4.7 |
| 2nd wave | 4126k | 10.63 | 0.946 | 10.5 |

It is notable that the *k*s for both 1st and 2nd wave are much larger than the previous waves in Taiwan because of the mitigated policy. The relaxations of measures are evaluated as the change of *k* from that in the Alpha surge which is about 0.05 leads to about 15 times enlargement of the number of daily confirmed case at a peak from Fig. 5 in Ref. [15]. It is concluded that “zero COVID” policy without lockdown sufficiently reduces the value of the probability *k*, when we compare the values for the Alpha surge. Also the value of *k* in the Omicron surge is comparable with that in Japan [14,15], where the monitoring method was not applied.

It is also worth noting that Taiwan did not change any major policy during the period of the Omicron surge. This fact is consistent with the natural suppression of infectious diseases due to voluntary behavioral changes assumed in the broken-link model.

### 3.3. The Omicron surge in Shanghai

In Shanghai, the Omicron surge occurred in early March is analyzed based on the broken-link model. In order to control the surge of COVID-19, Shanghai commenced the city-wide lockdown in late March, dividing the city into east and west districts (from 28th March for the east district and from 1st April for the west district). In each of the east and west districts, all residents were given two PCR testings at the beginning of lockdown. While those who tested positive were sent to a medical facility, those who had close contacts with a tested positive person were placed in an isolation facility. This is called the half-lockdown [23].

Subsequently, the full-lockdown [24–26] was commenced on 11th April, in which the entire city was managed in three areas as follows,

1. “blockade area” : The area was closed and one was not allowed to leave the house. An attendant visit them to supply foods or if necessary.
2. “controlled area” : Walks, etc. within a small area were allowed, but it was prohibited to gather or leave one’s area.
3. “precautionary area” : One could do anything only within the administrative ward.

These measures were important to see how effective severe behavioral restrictions on people had been against the spread of infectious diseases.

Fig. 3 (a) shows the daily confirmed cases including both symptomatic and asymptomatic cases in Shanghai from mid March to June 2022 and the corresponding K-values are given in Fig. 3 (b). These curves are plotted with the data reported by the China-CDC (Chinese Center for Disease Control and Prevention) [27]. It is worth mentioning that the epicurve has a peak just after the commencement of full-lockdown measures.

**Figure 3.**
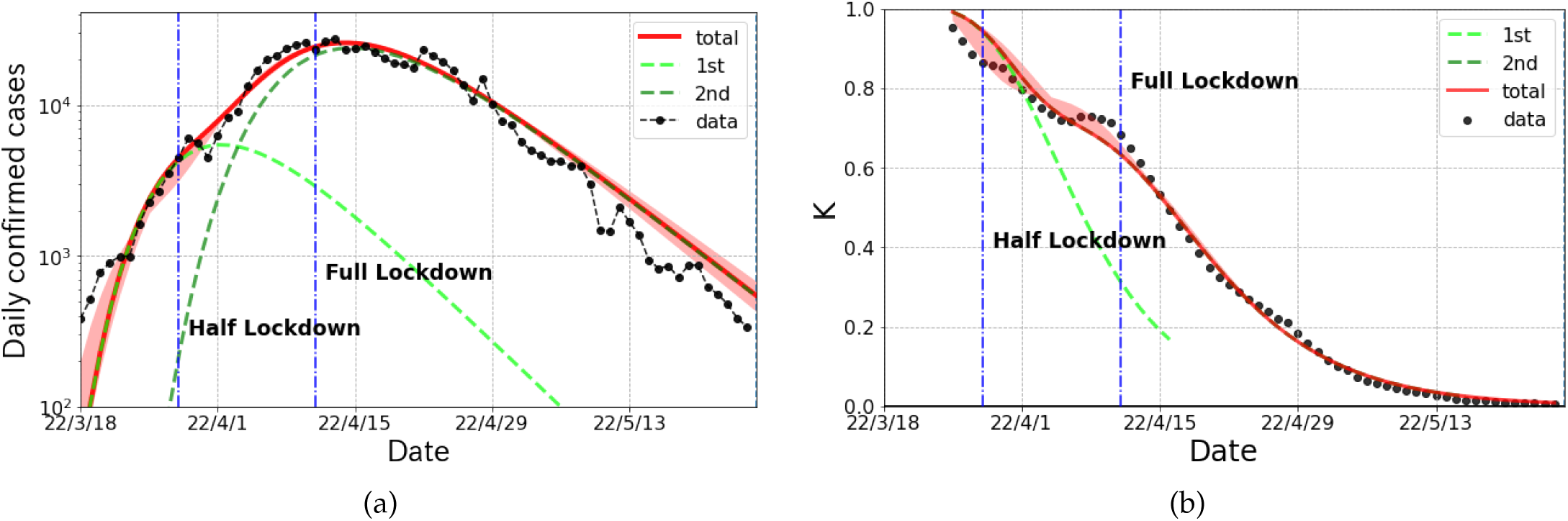
The logarithmic plot of the epicurve for the Omicron surge of COVID-19 in Shanghai from March to June 2022. (a) The daily confirmed cases and fit results (solid curve). The fit is performed with two partial waves denoted by dashed lines. (b) The observed data and fit results of the K-value. The red bands stand for systematic errors for the choice of fit range of the first and second partial wave.

This fact does not lead the conclusion that only the full-lockdown is meaningful to reduce the daily cases even the half-lockdown is comparatively strong measure.

The broken-link model revealed that the epicurve was consisted of two partial waves. As shown in Fig. 3 (a), the magnitude of 1st wave was much smaller than that of 2nd wave. The curve by the broken-link model nicely reproduce the epicurve and K-values in Shanghai. The 1st and 2nd wave are considered to be caused by BA.2 and by BA.2.2, respectively. The parameters of each partial wave are given in Table 3.

**Table 3.** The parameters of the Gompertz curves in the Omicron surge in Shanghai. The definition of the parameters is the same as in Table 1, but the reference date is 18th March 2022.

| partial wave | $N_\infty$ | $R_0$ | $k$ | Shift |
| --- | --- | --- | --- | --- |
| 1st wave | 98k | 6.89 | 0.865 | -0.1 |
| 2nd wave | 565k | 8.64 | 0.891 | 8.0 |

## 4. Discussion

We analyze the daily confirmed cases for the Alpha and the Omicron surge in Taiwan and the Omicron surge in Shanghai by using the broken-link model. These data are nicely described by the combination of partial wave components.

In Taiwan, though the lockdown method was not taken for both the Alpha and the Omicron surge periods, they changed from “zero COVID” policy [5] in period of the Alpha surge to the “pandemic management” policy [18–20] in period of the Omicron surge by mitigating the measures. The drastic policy change in Taiwan decreased the broken-link probability 1 - *k*, which cause about 15 times larger number of daily confirmed cases at a peak position.

We also find that the *k* is similar in magnitude under the same policy. This fact is helpful to predict the size of infectious diseases by the broken-link model.

Next, we discuss the Omicron surge in Shanghai. As shown in Fig. 3, the broken-link model nicely reproduces both daily confirmed cases and the K-values. From Table 3, both of these two waves have large basic reproduction numbers as *R*_0_ = 6.89 and 8.64, while both of *k*s are at most 0.89 which is quite smaller comparing with the other countries under the Omicron surge [15]. In spite of large *R*_0_s in both waves, the cumulative cases in period of the Omicron surge is only up to 2.4% of population because of small *k*. The *k* depends on a level of behavioral restrictions, political measures, immunity of human, etc. Of cause, it also depends on the strictness of lockdown measures but is considered to be far less sensitive under the stringent monitoring the activity of people under quarantine or isolation because whole epicurve can be reproduced with single *k* including before and after the commencement of lockdown. We rather have to take care of the risk on the commencement of lockdown since residents are converged for hoarding commodities against it. Such behavior of the population possibly enhances COVID-19 patients.

As a counterfactual hypothesis, we consider the 2nd partial wave in the Omicron surge in Shanghai by changing solely the link-connected probability to *k* = 0.920 which is the typical value in Japan [14,15]. The result of a counterfactual hypothesis shown in Fig. 4 indicates that the maximum number of infected persons per day is 23 times larger and the size of infected people is 30 times larger than the actual numbers. This can be considered as the effect of stringent behavioral restrictions against a COVID-19 spread.

**Figure 4.**
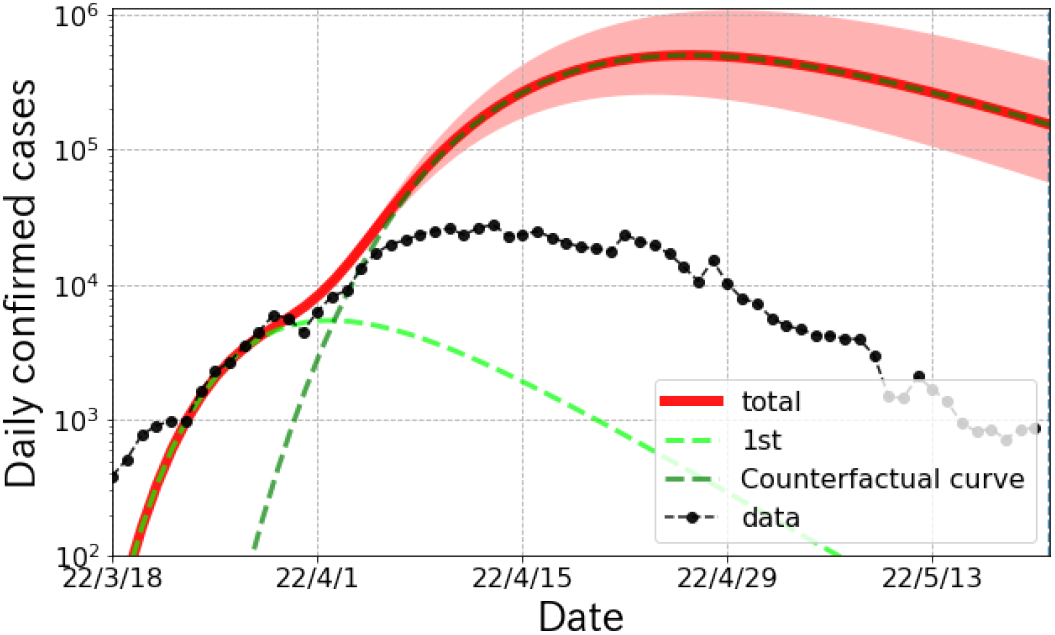
The logarithmic plot of the counterfactual hypothesis curve in the 2nd wave in Shanghai by changing the *k* from 0.891 to 0.920 which is typical value in Japan [14,15]. The red bands stand for the fluctuations with the ±0.05 changes in *k*.

It is interesting to compare the results of the Omicron surge in Taiwan as the mitigated behavioral restrictions and in Shanghai as the stringent behavioral restrictions with the similar population size. The size of cumulative cases and the link connection probability in Taiwan is 6 times and 5% larger than that in Shanghai, respectively. This difference is attributed to the mitigation behavioral restriction policy to recover the economical activity to the normal. Therefore, we have to think how to save the citizen’s live and economy from infectious diseases with the considerations of the effectiveness of behavioral restriction measures and the virulence of causative viruses.

## 5. Conclusions

We found that the link connection probability *k* defined in the broken-link model which mainly controls the size of cumulative cases is insensitive to lockdown measures if the activity of people under isolation or quarantine is strictly monitored. Although there is discussion that the behavioral monitoring of person may be an invasion of privacy, the stringent monitoring method are effective to suppressing the sarge of infectious disease. On the other hand, we found that the link connection probability *k* is not to be very small even if the strict lockdown is taken. It indicates that there are unavoidable infections or transmission from one’s intimate neighbors if infectious disease with high infectivity intrude one’s community.

## Data Availability

All data produced in the present work are contained in the manuscript

## Author Contributions

All authors contributed to the interpretation of the results obtained in this study and the final manuscript.

## Institutional Review Board Statement

Not applicable

## Informed Consent Statement

Not applicable

## Data Availability Statement

Not applicable

## Acknowledgments

We thank all the member of Division of Scientific Information and Public Policy (SiPP) at Center for Infectious Disease Education and Research (CiDER) Osaka University for useful discussions. This research was supported by The Nippon Foundation - Osaka University Project for Infectious Disease Prevention.

